# Episodic memory impairment in patients with repetitive head injury: Initial findings of the SNAP-CTE cohort study

**DOI:** 10.1101/2022.07.07.22277393

**Authors:** Ellen Erskine, Jennifer Batchelor, Michelle Maddren, Eamon Brown, Reidar Lystad, Rowena Mobbs

## Abstract

**Background:** Traumatic encephalopathy syndrome (TES) is considered a long-term, neurodegenerative consequence of repetitive head injury (RHI). This cohort study aimed to characterise the episodic memory profiles (specifically, immediate and delayed memory) of individuals with RHI history through neuropsychological assessment. Hypotheses included participants demonstrating reduced episodic memory functioning, and greater reductions in functioning observed with greater years of RHI exposure.

**Methods:** Neuropsychological assessment was conducted on 34 adults with ≥10 years of RHI exposure as a cohort study. Main outcome measures were auditory memory indices (AMI), immediate memory indices (AII), and auditory delayed memory indices (ADI). Other potential predictors of outcome variables (in addition to years of exposure duration) were also measured and factored into analysis so that they could be controlled for.

**Results:** AMI [*t* (33) = -2.4, *p* = .020), AII (*t* = -2.7, *p* = .012), and ADI (*t* = -2.7, *p* = .44) were all significantly below normative levels. AMI [*t* (33) = 4.1, *p* <.001), AII (*t* = 4.3, *p* <.001), and ADI (*t* = 3.7, *p* <.001) were also significantly below participants’ measured premorbid functioning. None of the comorbidities that were considered as possible confounding variables predicted the relationship of any outcome variables.

**Conclusions:** Previous research (1) indicated that immediate episodic memory (i.e., encoding) impairments do not appear to be associated with RHI, and our study provides evidence to the contrary. However, further research is required on larger sample sizes to further understand the relationship between RHI and encoding deficits in this complex population.

**What is already known on this topic?:** Chronic Traumatic Encephalopathy (CTE) identified at autopsy, has been loosely associated with a history of repetitive head injury (RHI) sustained in life, yet factors that account for symptoms such as defined in life as Traumatic Encephalopathy Syndrome (TES) which can include abnormal cognitive function, behavioural dysregulation and mood disturbances in this patient cohort are yet to be comprehensively investigated.

**What this study adds?:** This research is beneficial to the scientific community mainly because it contributes to the pre-existing body of literature on the neuropsychological profile of RHI. Previous research (1) has indicated that immediate episodic memory (i.e., encoding) impairments do not appear to be associated with RHI, and our study provides evidence to the contrary.

**How might this study affect research, practice, or policy?:** This research represents progress towards further discerning a neuropsychological profile of TES, thereby potentially aiding a better clinical diagnostic presentation of the disease, which can build on potential earlier diagnosis, prevention strategies and treatment pathways.

## Introduction

Concussion is recognised as a mild traumatic brain injury (TBI) and defined by American Academy of Neurology as a “clinical syndrome of biomechanically induced alteration of brain function typically affecting memory and orientation, which may involve loss of consciousness” (2). Sub-concussive head injury involves the transfer of mechanical energy to the brain with enough force to injure axonal or neuronal integrity and is considered below the threshold to elicit any signs of a concussion (3). Concussive and sub-concussive head injury most frequently occur within a sporting context, with over 54% of all concussions reported to be sporting-related (4). In sport, sub-concussive events are of particular concern as they tend to go unnoticed, and therefore players fail to be removed from play or medically evaluated (5). Athletes are also at considerably greater risk of repetitive head injury (RHI), with research indicating that athletes that sustain one concussion are statistically more likely to sustain future concussions (6). Moreover, largely because of lifestyle and post-injury factors, there is a high and varied prevalence of comorbidities associated with TBI and/or RHI and former athletes relative to the general population, including depression, post-traumatic stress disorder (PTSD), substance abuse, sleep apnoea, chronic pain, migraine, and stroke (7-9), subsequently making this a complex population to investigate.

In some individuals, a history of RHI has been associated with a later onset of progressive mood and cognitive impairment that correlates with radiological alterations and structural pathology at autopsy (10). This neurodegenerative disease, characterised by the accumulation of p-tau as neurofibrillary tangles within the brain (10), has been termed Chronic Traumatic Encephalopathy (CTE). The term TES, the terminology of preference in this research paper, has been coined in order to distinguish the clinical syndromes from the neuropathologically-defined CTE (11). The variety of clinical symptoms of TES was first associated with boxing in the 1920s, where abnormalities were described in nearly half of the long-term fighters, including dramatic personality disturbances, dysarthria, or Parkinson-like disturbances (12). In more recent years, TES has been associated with other contact sports, including American football (13), soccer (14), Australian rugby league (15), Australian rules football (16), and hockey (10). TES has also been recognised in individuals who have undergone non-sporting impacts, such as in epileptics and domestic abuse victims (17) as well as military veterans (18).

Currently, CTE can only be definitively diagnosed at autopsy through the identification of its characteristic neuropathology – however, varied clinical criteria for identifying TES in life have been proposed across the literature. Most recently, Katz et al. (11) developed consensus research diagnostic criteria for probable TES, with diagnosis requiring substantial exposure to RHIs, core clinical features of cognitive impairment (in episodic memory and/or executive functioning) and/or neurobehavioural dysregulation, a progressive course, and that the clinical features are not fully accounted for by any other neurologic, psychiatric, or medical conditions. Length of exposure appears to be a moderating factor in the development and severity of TES, with the greater number of years spent in a contact environment related to greater severity of disease burden (19). Duration played in American football players has been significantly associated with odds of TES at death, with odds increasing 30% every year, doubling every 2.6 years, and increasing by >10-fold every nine years. A threshold of approximately 11 years of play was recommended in order to maximise the sensitivity and specificity together of developing TES (19). On the other hand, cognitive reserve (typically measured through levels of intelligence and/or educational and occupational attainment) may serve as a protective factor, whereby individuals with higher reserve can sustain greater brain damage before demonstrating functional deficit (20). A preliminary examination conducted by Alosco et al. (21) found a positive correlation between cognitive reserve (as indicated by occupational attainment) and later symptom onset of TES (as measured by retrospective next-of-kin interviews) in a sample of 25 former professional American football players with neuropathically-confirmed TES. While this was a preliminary cohort study with limited generalisability and a reliance on potentially biased self-report measures, the results suggest that higher cognitive reserve in individuals with TES is likely to attenuate and delay onset of cognitive impairment.

Neuropsychological assessment is critical in gaining a detailed understanding of the clinical profile of TES, as it can identify subtle patterns of cognitive and behavioural impairments which may not be apparent on other measures, such as brain imaging or self-report (22). While there has been little exploration of TES patient performance on neuropsychological assessment, there is extensive data on neuropsychological performance following TBI, which can serve as a reasonable foundation on which to predict the cognitive functions most likely to be affected in TES. Episodic memory has been established as one of the functions that is particularly impaired in TBI (23). A meta-analysis of the effects of moderate-to-severe TBI on episodic memory (23) found that verbal memory, as measured on neuropsychological assessment by recall of a word list (e.g., the California Verbal Learning Test, Second Edition; CVLT-II) and/or a story (e.g., Logical Memory from the Wechsler Memory Scale, Fourth Edition; WMS-IV), showed the largest effect size (*g* = .94). It was concluded that verbal recall measures demonstrate higher sensitivity to TBI effects on memory processes and should be routinely implemented in memory testing of TBI populations. Episodic memory impairments have also been implicated in TES (11, 17), and would therefore be important to measure in the neuropsychological assessment of patients with possible TES.

While there is evidence for an association between history of RHI and chronic cognitive and behavioural impairment that can be observed in TES, there is insufficient data to confirm a causal link. This is partly attributed to the fact that individuals with a history of RHI have only been studied retrospectively following their deaths, and therefore clinical phenotypes cannot be reliably linked with autopsy-confirmed TES (24). While consensus diagnostic criteria have been recently developed to aid research into the neurological pathology following concussion and repeated mTBI (11), their utility as clinical diagnostic criteria for TES remains to be determined. Moreover, as previously mentioned, the population of individuals with TBI and/or RHI are also more likely to be diagnosed with comorbidities unrelated to TES compared to the general population (7-9). It has therefore been argued that the symptoms associated with TES may be related to other such contributors (8). Accordingly, there remains a degree of scepticism over TES, and whether it exists as a discrete clinical syndrome (25). There is therefore a clear need for methods to identify TES during life. Not only would this assist in validating TES as a distinctive disease but more importantly, would aid early diagnosis and form the basis for clinical trials in the treatment and prevention of the disease.

The current research is a part of a larger, longitudinal project which aims to characterise the clinical, neuropsychological, neurophysiological, neuro-ophthalmological and neuroradiological changes in an Australian population with a history of RHI by gathering these different kinds of data from participants every two years until death. The aim of the current study was to characterise the episodic memory profiles (and specifically, immediate and delayed memory) of individuals with a history of RHI at self-identified risk of TES through neuropsychological assessment. The following hypotheses were tested:

1. Relative to both normative data and participant premorbid functioning, participants will demonstrate reduced functioning in episodic memory.
2. A greater number of years spent in an environment associated with RHI exposure will be associated with greater reductions in episodic memory functioning.

## Method

### Participants

Participants included 34 adults (31 males, 3 females) between the ages of 24 and 64 years that were self-identified as at risk of sequelae of RHI. All participants were initially seen by a neurologist who assessed their suitability for inclusion in the study. Exclusion criteria included: i) individuals aged <18 years, ii) aged ≥65 years, iii) <10 years of RHI exposure (derived from the threshold of 11 years established by Mez et al. (19)), iv) performed below cut-scores on an effort measure, v) previous neurosurgery, birth trauma or serious malignancy, and vi) a diagnosis of a neurodegenerative disorder other than TES.

Patients and Public Involvement was not included in this study, as the study was using set Neuropsychological tests which have been independently validated. This study simply shows the results of these tests which is inconsistent with other findings in the literature. Further studies that will expand on this study, may include more comprehensive patient and public involvement.

Demographic information regarding the participants and their RHI history is included in Table 1.

**Table 1.** Demographics and exposure type associated with concussion data (n = 34)

Characteristic
|  |  |
| --- | --- |
| Age (years), M (SD) | 49.2 (10.5) |
| Gender, n of males (%) | 31 (91.2) |
| Level of education, n (%) ($M = 13.3$, $SD = 2.8$) | |
| <10 years | 2 (5.9) |
| School certificate (10 years) | 5 (14.7) |
| Higher school certificate (12 years) | 10 (29.4) |
| 13-15 years | 7 (20.6) |
| >15 years | 10 (29.4) |
| Length of RHI exposure (years), M (SD) | 22.3 (8.1) |
| Length of RHI exposure for $\geq 2$ exposure types (years), M (SD) | 21.6 (7.2) |
| 1 Exposure Type ($n = 18$, 52.9%) | 23.0 (9.0) |
| 2 Exposure Types ($n = 10$, 29.4%) | 23.8 (7.5) |
| 3 Exposure Types ($n = 3$, 8.8%) | 14.0 (4.6) |
| 4 Exposure Types ($n = 3$, 8.8%) | 21.7 (2.9) |
| Exposure type, n (%)* |  |
| Rugby Union | 14 (41.2) |
| Rugby League | 12 (35.3) |
| Boxing | 6 (17.6) |
| Bike riding | 4 (11.8) |
| Soccer | 4 (11.8) |
| Australia Football League | 2 (5.9) |
| Basketball | 2 (5.9) |
| Domestic violence | 2 (5.9) |
| Falls | 2 (5.9) |
| Horse racing | 2 (5.9) |
| Judo | 2 (5.9) |
| Karate | 2 (5.9) |
| Military | 2 (5.9) |
| Water skiing | 2 (5.9) |
| Parachuting | 1 (2.9) |
| Softball | 1 (2.9) |
| Wrestling | 1 (2.9) |

Note: M = mean, SD = standard deviation, n = sample size, RHI = repetitive head injuries
\*16 participants were associated with $\geq 2$ exposure types (47.1%)

### Procedure

All neuropsychological assessments were conducted by registered clinical neuropsychologists or clinical neuropsychological post-graduate students using standardised administration as outlined in test manuals. Of these, 24 were conducted via standard face-to-face assessment and 10 were conducted online via a telehealth program due to COVID-19 related restrictions. Assessment began with an initial semi-structured clinical interview to ascertain the participant’s RHI exposure history, and psychological, medical, and educational history.

#### Confounding variables

Possible confounding variables were factored into analysis so that they could be controlled for. These included: i) cognitive reserve, as measured by years of education, with a greater number of years of education likely predicting higher cognitive reserve and thus higher episodic memory functioning, ii) premorbid functioning, as measured through neuropsychological measures, with higher scores demonstrating higher premorbid functioning and thus predicting higher episodic memory functioning, iii) psychological distress, as measured through a self-report questionnaire, with higher levels of psychological distress likely predicting lower episodic memory functioning, and iv) any comorbidities that were prevalent in our sample.

Comorbidities identified in participants are detailed in Table 2. Migraine and alcohol-use disorder were diagnosed by the neurologist in accordance with the International Classification of Headache Disorders (ICHD-3; (26)) and Diagnostic and Statistical Manual of Mental Disorder (DSM-5; (27)) criteria respectively. Sleep apnoea was previously diagnosed by a sleep/respiratory physician. Prior diagnoses of PTSD, chronic pain, and neurological disorders as well as past drug-use were noted during clinical interview and on participant’s medical records.

**Table 2.**
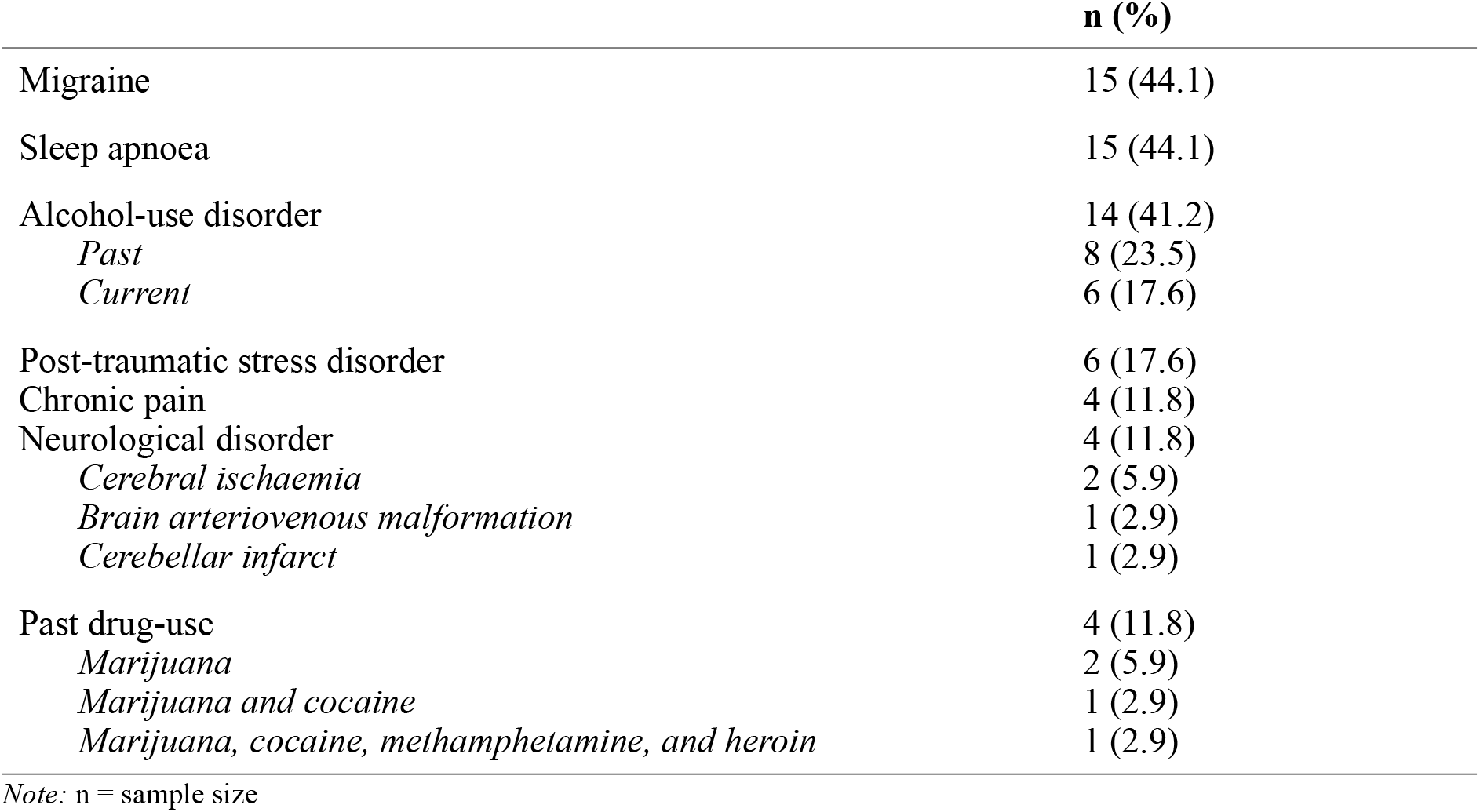
Comorbidities

|  | **n (%)** |
| --- | --- |
| Migraine | 15 (44.1) |
| Sleep apnoea | 15 (44.1) |
| Alcohol-use disorder | 14 (41.2) |
| *Past* | 8 (23.5) |
| *Current* | 6 (17.6) |
| Post-traumatic stress disorder | 6 (17.6) |
| Chronic pain | 4 (11.8) |
| Neurological disorder | 4 (11.8) |
| *Cerebral ischaemia* | 2 (5.9) |
| *Brain arteriovenous malformation* | 1 (2.9) |
| *Cerebellar infarct* | 1 (2.9) |
| Past drug-use | 4 (11.8) |
| *Marijuana* | 2 (5.9) |
| *Marijuana and cocaine* | 1 (2.9) |
| *Marijuana, cocaine, methamphetamine, and heroin* | 1 (2.9) |

Note: n = sample size

### Measures

Standardised cognitive measures were selected based on previous research indicating their sensitivity to TBI-related episodic memory impairment. All participants were assessed for suboptimal effort using an embedded measure of Reliable Digit Span from the Wechsler Adult Intelligence Scale, Fourth Edition (WAIS-IV; (28)), utilising a cut-off score of ≤6 (29). Due to the COVID-19-related lockdowns, solely verbal/auditory-based tests were utilised to allow for ease-of-administration via telehealth.

#### Premorbid functioning

The three subtests of the WAIS-IV (28) that comprise the Verbal Comprehension Index (VCI; i.e., Similarities, Vocabulary, and Information) were administered to estimate premorbid functioning. The VCI is considered a good predictor of overall intelligence that is resilient to the effects of brain impairment relative to other cognitive domains (30). It has a high average split-half reliability and test-rest reliability (both .96; (31)). A measure of premorbid functioning was necessary as a basis of comparison with the outcome measures, whereby inferences can be drawn regarding whether or not the outcome measures reflect some decline from premorbid functioning (32). Raw data was analysed using the Advanced Clinical Solutions (ACS; (33)) software package to produce age-normed VCI composite scores (*M =* 100, *SD* = 15).

#### Immediate and delayed memory

Memory testing included the four subtests of the WMS-IV (34) that comprise the Auditory Memory Index (AMI; i.e., Logical Memory I and II and Verbal Paired Associates I and II). Raw data was again analysed using the ACS software package to produce age-normed AMI (an overall measure of auditory memory), Auditory Immediate Index (AII; a measure of the ability to encode or commit novel verbal material to memory), and Auditory Delayed Index (ADI; a measure of the ability to retain material over time) (*M =* 100, *SD* = 15). AMI has a high average internal consistency and test-rest reliability (.95 and .81, respectively; (35)). AII and ADI have moderate to high internal reliabilities across most age groups (0.80s to 0.90s) and a moderate to high test-retest reliability (.50 to .82; (36)).

#### Psychological distress

The Depression, Anxiety and Stress Scale (DASS-21; (37)) was utilised as a measure of participant’s current level of psychological distress. For ease of analysis, subscale scores were combined to yield a total score ranging from 0 to 63, with higher scores indicating higher levels of psychological distress. The literature indicates that the three scales can be incorporated to form one ‘general distress’ factor, with the total DASS-21 score having a very high internal consistency of .93 (38).

## Results

Descriptive statistics of all neuropsychological measures are described in Table 3.

**Table 3.** Descriptive statistics of neuropsychological measures

| Measure | Score type | *n* | *M* | *SD* |
| --- | --- | --- | --- | --- |
| VCI | Composite | 34 | 103.8 | 12.2 |
| AMI | Composite | 34 | 92.7 | 17.4 |
| AII | Composite | 34 | 92.0 | 17.5 |
| ADI | Composite | 34 | 92.8 | 17.5 |
| DASS-21 | Raw | 34 | 18.7 | 14.4 |

*Note:* *M* = mean; *SD* = standard deviation; *n* = sample size; VCI = Verbal Comprehension Index; AMI = Auditory Memory Index; AII = Auditory Immediate; ADI = Auditory Delayed Index; DASS-21 = Depression, Anxiety and Stress Scale – 21 items.
Composite Normative Scores: $M = 100$ , $SD = 15$ .

As determined by Shapiro-Wilk tests, all outcome variables were normally distributed (all *p*-values ≥.25). Using one-sample *t*-tests, composite scores obtained on indices derived from the WMS-IV (AMI, AII, and ADI) were compared to a normative score of *M* = 100 (*SD* = 15) as determined by Wechsler (28). Results are detailed in Table 4. There was a statistically significant difference for AMI, AII, and ADI, with those in the sample performing significantly below normative levels on all measures (all *p*-values ≤.023).

**Table 4.** One Sample t-test Results (Compared with a Normative Score)

| Index | *M* difference | *t* | *df* | *p* | *d*<sup>a</sup> |
| --- | --- | --- | --- | --- | --- |
| AMI | -7.3 | -2.4 | 33 | .020* | .45 |
| AII | -8.0 | -2.7 | 33 | .012* | .49 |
| ADI | -7.2 | -2.4 | 33 | .023* | .44 |

Note: M = mean; df = degrees of freedom; n = sample size; AMI = Auditory Memory Index; AII = Auditory Immediate Index; ADI = Auditory Delayed Index.
Cohen's $d^a$ effect sizes: .20 = small effect; .50 = medium effect; .80 = large effect
\* $p < .05$

Using paired-sample *t*-tests, AMI, AII, and ADI scores were then compared to VCI (premorbid functioning) scores. Results are detailed in Table 5. There was a statistically significant difference between AMI, AII, and ADI and VCI, with AMI, AII and ADI being significantly lower than VCI (all *p*-values <.001).

**Table 5.** Paired Sample t-test Results when Compared with VCI Scores

| Index | $M$ difference | $t$ | $df$ | $p$ |
| --- | --- | --- | --- | --- |
| AMI | -11.1 | 4.1 | 33 | $< .001^*$ |
| AII | -11.8 | 4.3 | 33 | $< .001^*$ |
| ADI | -11.0 | 3.7 | 33 | $< .001^*$ |

Note: M = mean; df = degrees of freedom; VCI = Verbal Comprehension Index; AMI = Auditory Memory Index; AII = Auditory Immediate Index; ADI = Auditory Delayed Index.
\* $p < .01$

### Forward Selection Linear Regression

Forward selection linear regression analyses were used to identify potential predictors of all outcome variables from the following candidate variables: VCI (premorbid functioning), years of education, exposure duration, DASS-21 scores (psychological distress), alcohol-use disorder (AUD), sleep apnoea, and migraine (with the latter three analysed as binary variables coded 1 = Yes, 2 = No). Age was not included in analyses, as all scores were already corrected for age when originally converted to an aged-normed standardised score. Gender was also not included given the very skewed distribution (91.2% males), nor was PTSD, chronic pain, neurological disorder, and past drug-use given their low sample size relative to other comorbidities (*n* ≤ 6).

Firstly, univariate regression analyses were conducted on each of the outcome variables to determine their relationship with each of the possible predictors, and thus the need to control for any potential predictors in the final model. *P*-values of these results are detailed in Table 6. A more lenient significance threshold of *p* ≤.25 was utilised to set a limit on each potential predictor’s inclusion in the final model, as more traditional cut-off levels such as .05 can fail in identifying important predictors (39). On this basis, VCI, years of education, AUD, and sleep apnoea were included in the model examining possible predictors of AMI. The AII model included VCI, years of education, DASS-21 scores, AUD, and sleep apnoea. The ADI model included VCI, years of education, and sleep apnoea.

**Table 6.**
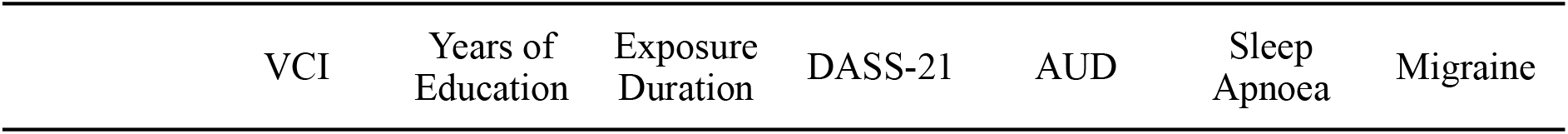
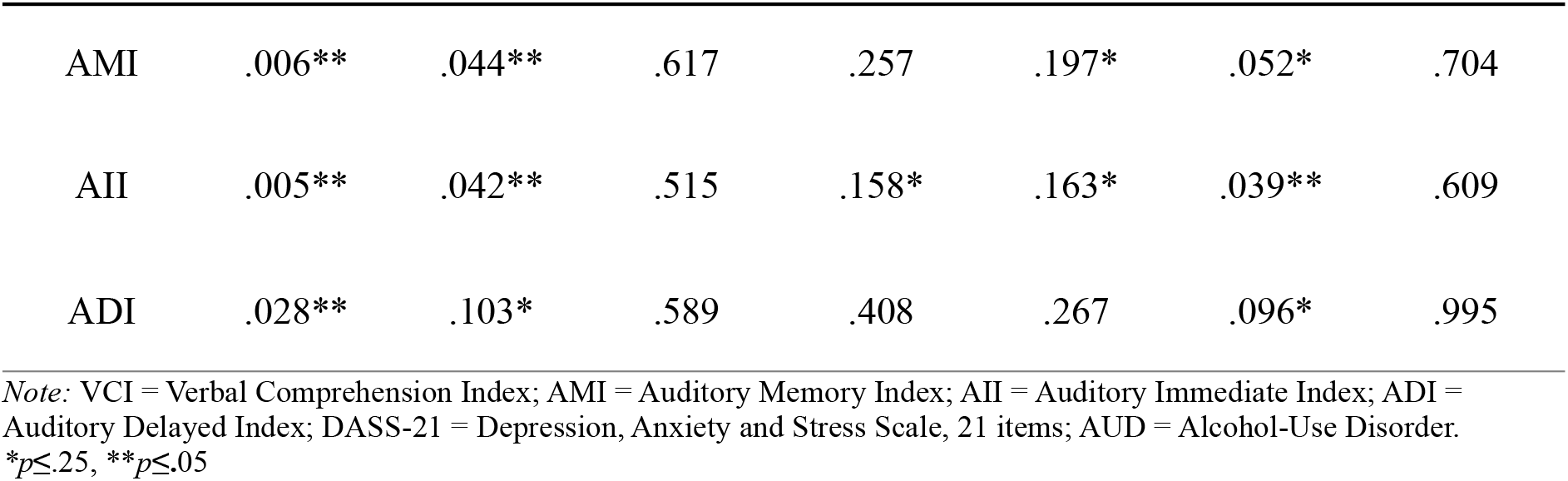
Univariate Regression Analyses p-values

Secondly, multivariate regression analyses were conducted on each of the models. VCI, years of education, alcohol-use disorder, and sleep apnoea were significant overall predictors of AMI, *F*(4,29) = 3.2, *p* = .026, with an *R*^*2*^ of .3. However, no predictors significantly contributed to the model in isolation. VCI, years of education, DASS-21 scores, alcohol-use disorder, and sleep apnoea were significant overall predictors of AII, *F*(5,28) = 2.97, *p* = .036, with an *R*^*2*^ of .3. Again, no predictors significantly contributed to the model in isolation. VCI, years of education, and sleep apnoea were not significant overall predictors of ADI, *F*(3,30) = 2,4, *p* = .086, with an *R*^*2*^ of .23, and no variables significantly contributed to the model in isolation.

## Discussion

The aim of the current cohort study was to characterise the episodic memory profiles (specifically, immediate and delayed memory) of individuals with a history of RHI at self-identified risk of TES through neuropsychological assessment. It was firstly hypothesised that participants would demonstrate reduced functioning in episodic memory. This hypothesis was supported: As a group, participants demonstrated significantly reduced functioning across all measures of episodic memory relative both to normative data and participant’s premorbid functioning. This finding is consistent with the TBI literature (23) and research based on retrospective reviews of neuropathologically-confirmed CTE meeting the criteria of TES (11, 17) indicating impairment in episodic memory. Here, it is most likely that the dysfunction primarily lies in immediate auditory memory, i.e., encoding (intake and organization) of information. As observed in studies investigating memory deficits in Alzheimer’s disease, if participants were demonstrating particular difficulties with delayed memory (storage and/or retrieval of information), we would expect reduced delayed memory performance relative to immediate memory performance (40), which was not the case. This is in contrast to previous research by Wright et al. (1) comparing the memory profiles of retired NFL players with a history of RHI against individuals with single-impact TBI (relative to controls) on a list-learning task (the CVLT-II). These findings indicated that RHI is primarily associated with storage and retrieval deficits, but not encoding deficits (1). This discrepancy in findings may be attributable to some of the methodological limitations of Wright et al.’s study (1), such as not controlling for all potential confounding variables and participants’ premorbid functioning.

We also hypothesised that higher exposure duration (i.e., years spent in an RHI environment) would be associated with greater reductions in episodic memory functioning (19). However, contrary to this prediction, exposure duration did not significantly predict scores on any outcome measures. It is possible that, in this sample, exposure duration did not sufficiently encapsulate the quantity and severity of participant’s RHI exposure because of the significant variability in types of exposure environments across participants, each likely having very different head impact severity and frequencies. However, measuring the number of head injuries sustained was not a feasible alternative, as it can be almost impossible to measure the exact number of RHIs obtained across a whole sporting career due to the unreliability of self-report measures and the difficulty in recognising sub-concussive head impacts.

Finally, we anticipated that there would be protective (i.e., cognitive reserve and premorbid functioning) and risk factors (i.e., exposure duration, psychological distress, and comorbidities that were identified as prevalent in our sample) potentially influencing the significance and direction of the outcome variables. However, none of these factors appeared to significantly predict the relationship of any outcome variable when factored into the final model. This therefore supports the finding that impairments in episodic memory are related to RHI rather than other such potential contributors.

There are several limitations of this cohort study that need to be addressed. Firstly, a relatively small sample size limits the statistical power and generalisability of our research to the wider population. Given the disproportionate number of males to females, it also needs to be recognised that conclusions drawn from this study did not control for differences in sex, with effect sizes most likely being favourably biased towards male experiences of RHI sequelae and TES. Furthermore, participants were selected only from among those already experiencing cognitive and/or behavioural complaints, thus potentially biasing the sample and limiting the applicability of the findings. While we attempted to minimise the impact of this selection bias by screening participants for response validity and exaggeration of cognitive impairment, cognitive impairment could still be overrepresented in our sample. Additionally, outcome variables were compared to normative data rather than an age-matched control group. There were also several comorbidities that were not controlled for in the study, including PTSD, chronic pain, neurological disorder other than TBI, and past drug-use, which was justified due to the very small occurrence of these comorbidities within the cohort. Finally, while a proportion of participants have consented to donate their brains after death to an active autopsy program, recruitment was not restricted to these individuals. Therefore, there will not be a way to definitively correlate those participants’ clinical profiles to a diagnosis of CTE. To address these limitations, future research conducted with a greater time span and sample size should aim to recruit a proportional number of females and minimise methodological issues that may have arisen as a result of having such a varied sample. For example, future research could consider the recruitment of only former football players with a similar number of years of RHI exposure. Additionally, to further extend this research, future research could also consider implementing a greater number of neuropsychological measures assessing additional cognitive domains such as executive functioning.

With regard to the strengths of the current study, this firstly includes possible variance accounted for by age being effectively controlled for through the use of age-normed data. We also attempted to control for potential confounding variables including cognitive reserve, premorbid functioning, psychological distress, and comorbidities prevalent within our sample. The inclusion of premorbid functioning as a basis of comparison with outcome variables also allowed for the consideration of whether the obtained scores reflected some decline from participant premorbid levels, rather than solely basing comparisons on normative data. Finally, we utilised an effort measure to control for exaggerated impairment or insufficient effort.

## Conclusion

This research is beneficial to the scientific community mainly because it contributes to the pre-existing body of literature on the neuropsychological profile of RHI. Previous research (1) has indicated that encoding deficits do not appear to be associated with RHI, and our cohort study provides evidence to the contrary. Most importantly, this research represents progress towards further discerning a neuropsychological profile of TES, thereby aiding in the opportunity for early diagnosis, treatment, and prevention of the disease.

## Data Availability

All data produced in the present study are available upon reasonable request to the authors.

